# Clinical validation of point-of-care SARS-COV-2 BD Veritor antigen test by a single throat swab for rapid COVID-19 status on hospital patients predominantly without overt COVID symptoms

**DOI:** 10.1101/2021.04.12.21255299

**Authors:** Jesper Bonde, Ditte Ejegod, Helle Pedersen, Birgitte Smith, Dina Cortes, Cæcilie Leding, Thorbjørn Thomsen, Thomas Benfield, Uffe Vest Schnieder, Jens Tingleff, Marc Arbyn, Gorm Lisby, Ove Andersen

**Author notes:** Corresponding author: Jesper Bonde [ ].

## Abstract

**BACKGROUND:** Fast identification of severe acute respiratory syndrome Coronavirus-2 (SARS-CoV-2) infected individuals is a strategically vital task to ensure correct management and quarantine. Rapid antigen test could be a supplement to the standard-of-care Nucleic Acid Amplification Test (NAAT). The aim of this study was to determine the accuracy of the BD Veritor SARS-CoV-2 antigen test as a screening instrument in a hospital setting.

**METHODS:** A cohort of prospective samples were collected from hospital staff and patients at the Emergency, Infectious Diseases and Pediatrics and Adolescent Medicine departments at Hvidovre Hospital. All samples were collected using oropharyngeal swabs, and BD Veritor Antigen test results were paired with routine NAAT test results. Sensitivity, specificity, positive and negative predictive values of the antigen test were calculated using NAAT as reference.

**RESULTS:** Overall, 809 samples from 674 individuals were included (average age 45 years, range 0-98 years). Among all samples, 8% were SARS-CoV-2 positive by NAAT testing and 5.3% by BD Veritor. The sensitivity of the antigen test was 63.1% and specificity 99.7%. The positive predictive value was 95.3%. False-positive rate was 4%. The cycle threshold value was significantly higher among individuals with false negative antigen tests compared to true positives.

**CONCLUSION:** The sensitivity, specificity and positive predictive values show that the BD Veritor antigen test from oropharyngeal collected specimens performs well. Antigen testing may be a supplement, but not substitute, to NAAT testing as the primary diagnostic modality in hospital settings where fast turnaround test results may assist in decisions regarding isolation and quarantine.

## Introduction

Fast and easy-to-perform diagnostic methods is of high priority to curb the Coronavirus Disease 2019 (COVID-19) pandemic. As the pandemic enters its 2^nd^ year, the political focus in many countries including Denmark is on easy test accessibility, fast time-to-result, accurate tests and cost-efficiency. Even when vaccination is fully rolled-out, it is anticipated that severe acute respiratory syndrome Coronavirus-2 (SARS-CoV-2) testing will remain a pivotal public health measure the coming years. The reference standard for SARS-CoV-2 testing is Nucleic Acid Amplification Test (NAAT) targeting the RNA genome of SARS-CoV-2 in upper-respiratory specimens and performed in centralized laboratory settings ^1,2^. NAAT systems deliver test-to-result in 2 to 6 hours depending on circumstances and this time-to-result has obtained widespread acceptance as a premise of protecting public health by identifying infected individuals and referring them into quarantine. However, many SARS-COV-2 test situations benefit from point-of-care testing (POCT) with fast time-to-result.

In a hospital setting, rapid identification of SARS-CoV-2 infected patients is a strategically vital task to ensure correct clinical management, precaution management, isolation or quarantine of SARS-CoV-2 positive patients. SARS-CoV-2 carriers pose a potential risk to healthcare professionals who are in close contact with the patient during examinations or treatments for non-COVID-19 as well as COVID-19-related conditions, and constitute a risk to other patients in e.g. common waiting areas and shared hospital rooms in high through-put Emergency Departments were a flow-culture is prominent^3^.

Antigen tests are immunoassays that detect a specific viral antigen. Rapid SARS-CoV-2 antigen detection (RAD) tests can be performed as a POCT, are easy to administer, and the results are typically available within 15-20 minutes. Combined with the solid political focus, especially on time-to-result and scalability, it has resulted in the implementation of SARS-CoV-2 antigen POCT in public and private test settings. The RAD test market is booming as the SARS-CoV-2 pandemic unfolds^4,5^, but qualitative data on their performance is limited and biased by test cohort characteristics^6^. SARS-CoV-2 antigen POCT is a promising instrument to optimize SARS-CoV-2 control in conjuncture with NAAT test regimens, but principal concerns regarding antigen test performance include low sensitivity, reproducibility, and low positive predictive value, all depending on specific circumstances^6,7^.

This study aimed to conduct a clinical performance validation of the BD Veritor antigen Rapid test (Veritor) using oropharyngeal swabs from hospital staff and patients (inpatients and outpatients). The majority of tested were without COVID-19 symptoms tested as a routine screening.

## Material and Methods

### Study cohort and Sample collection

The study was performed at Amager-Hvidovre Hospital (AHH), Copenhagen, Denmark, between December 2020 and March 2021.

A cohort of prospective collected samples from hospital staff and patients with unknown SARS-CoV-2 status arriving at the Department of Pediatrics and Adolescent Medicine, or the Emergency Department. The cohort was enriched with a smaller cohort of patients with known, recently NAAT confirmed SARS-CoV-2 positivity from the Department of Infectious Diseases to increase the number of positive specimens.

All samples were obtained by oropharyngeal swab (throat swab) in accordance with the Danish National Health Authority guidance for testing of SARS-CoV2. Similarly, a dedicated throat swab was used for the Veritor antigen test (FLOQSwab, Copan Diagnostics, Italy). All patients were handled according to guidelines as per their SARS-CoV-2 NAAT status. The Veritor result did not affect the management.

### BD VERITOR Ag Rapid test and reference test systems

The “*BD Veritor System for Rapid Detection of SARS-CoV-2*” is a chromatographic lateral flow immunoassay for the qualitative detection of nucleocapsid antigens in a respiratory specimen. The dedicated Veritor throat swab was resuspended for 15’sec in the provided test tube containing a virus transport medium that neutralizes SARS-CoV-2 virus to a non-infectious level^8^. The test cassette incubates for 15 minutes in the BD Veritor analyzer (VA) and the result is reported by the VA. Patient ID was scanned into the VA ensuring safe patient ID identification. The manufacturer reports a test specificity of 100% and a sensitivity of 84% compared to qRT-PCR as a reference standard during the first 5 days after disease onset for use on superficial nasal specimens (Manufacturers insert).

SARS-CoV-2 NAAT testing was part of the routine diagnostic for SARS-CoV-2 testing performed at either the Department of Clinical Microbiology, AHH or Testcenter Danmark, Statens Serum Institute, Copenhagen. Three hospital SARS-COV-2 NAAT test systems were in routine operation during the data collection period: Cobas FLOW, BGI, and Hologic Panther Aptima^®^ SARS-CoV-2 Assay. For Cobas FLOW, oropharyngeal samples were tested by RT-PCR targeting the E-gene applying the E-Sarbeco primers and probe^*9*^ and adapted to TaqMan Fast Virus 1-step master mix and LightCycler 480^*10*^. BGI and Hologic testing were according to the manufacturer’s specifications. Interpretation of NAAT results were by a commercial FDA approved NAAT with automatic calling of positive or negative results or by PCR with manual inspection of the amplification curve for all positive samples. Only positive PCR samples with an unambiguous amplification curve were registered as positive. Any of these three NAAT assays were used as reference standard for the evaluation the accuracy of Veritor.

### Data analysis

Sensitivity, specificity, Positive Predictive Value (PPV), Negative Predictive Value (NPV), false positive rates were calculated using NAAT as reference. A box plot depicting the Cycle Threshold (Ct) scores between concordant and discordant PCR-to-Veritor samples (limited to Cobas FLOW and BGI results) were analyzed in GraphPad Prism version 9.

### Statistical power

The cohort size was determined on >80% power to detect a 5% difference in sensitivity between PCR and the antigen POCT. A SARS-CoV-2 positivity rate of <2% with an increasing trend was assumed. Based on the above, at least 773 paired samples were required.

### Ethical approval

Unselected samples from in- and outpatients and staff constitute pilot implementation with embedded diagnostic quality development (Ethics Committee of the Capital Region of Denmark, record H-20080005). The selected SARS-CoV-2 positive patients’ samples were collected through the out-patient research team (H-20028292).

## Results

In total, 971 Veritor samples were collected from 779 persons (Figure 1). Samples with a Veritor test (index) and a reference NAAT test result within ±5 days were included. Twelve samples were included with a time difference between index and reference test >5 ≤10 days based on the premise that the paired reference was the only available and that results were concordant. Ninety-one samples were excluded due to no matching reference test. Seven samples were excluded due to incorrect personal identifiers. Forty-nine samples were excluded as multiple samples collected from the same subject within a 24 hours window. Eleven samples were excluded due to a history of prolonged SARS-CoV-2 positivity rendering the patient irrelevant for antigen test. Four samples were excluded based on insufficient sampling material. Seventy-two individuals were tested twice or more during the study period (11%, Figure 1). The Department of Pediatrics and Adolescent Medicine provided 49 samples from in- and outpatients (average age; 3.8 years, range 0-25 years). The Emergency Department provided 731 samples (average age 48 years, range 10-98 years) from patients and staff seen at the Corona Assessment Clinic or in The Emergency Department. The Department of Infectious Diseases provided 29 samples from individuals visited at home. These individuals had a recent positive NAAT test for SARS-CoV-2. In total, paired test results from 809 samples in 674 individuals were included (average age 45 years, range 0-98 years).

**Figure 1.**
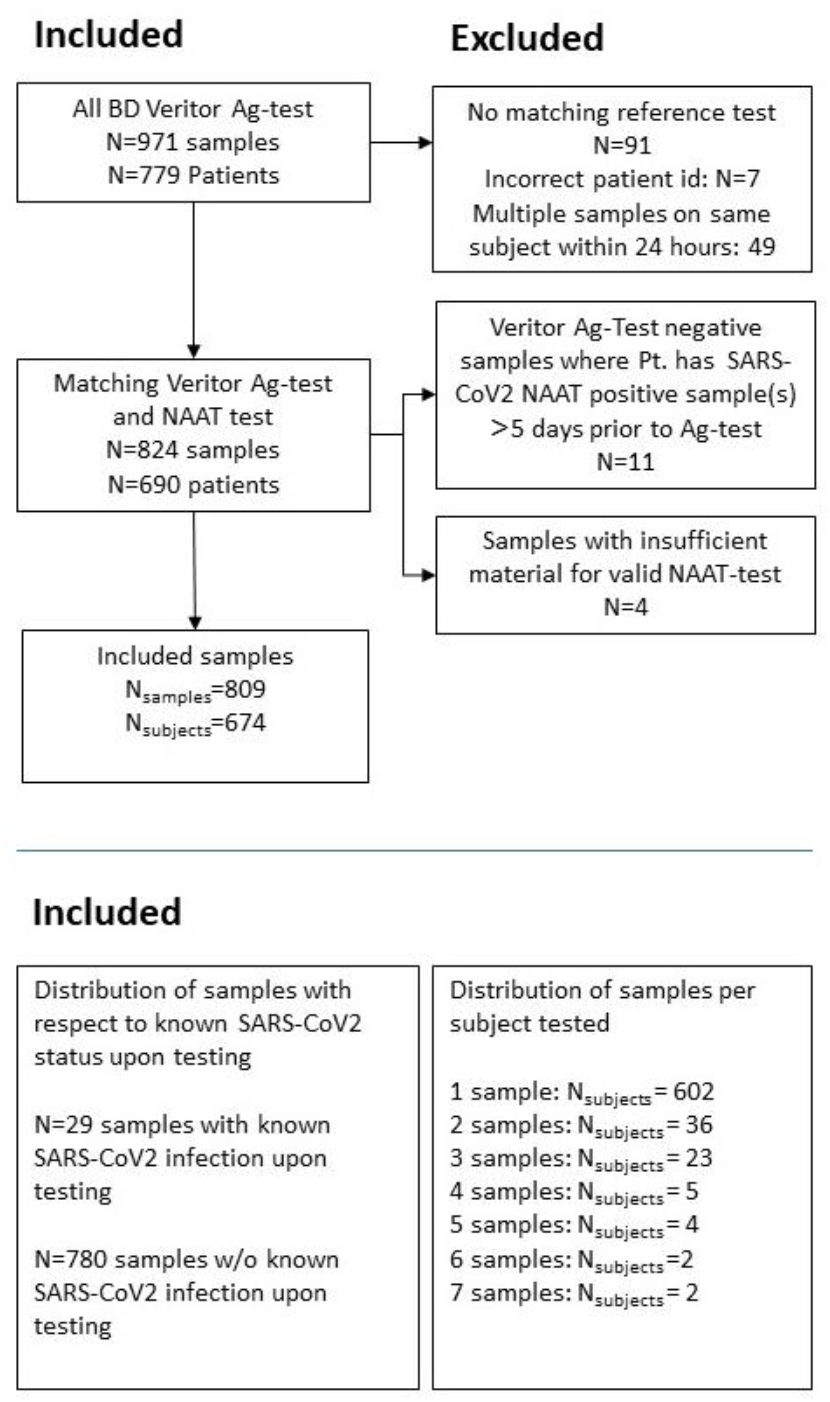
Flow chart describing inclusion and exclusion of samples and patients/persons in the study cohort. Below the line, a detailed description of the distribution between patients with known SARS-CoV-2 positivity and persons w/o known SARS-CoV-2 status at time as testing, and a listing of the number of samples per person included in the cohort.

PPV for the overall population, patients with known and recent NAAT confirmed SARS-CoV-2, and for individuals with unknown SARS-CoV-2 status were 95.3%, 94.4% and 96.0%, respectively. NPV values for overall and individuals with unknown SARS-CoV-2 status were 96.9% and 97.6%, respectively. The Veritor false-positive rate amongst individuals with unknown SARS-CoV-2 status was 4.0% (1/25).

The prevalence of SARS-CoV-2 as determined by NAAT was 8.0% (65/809) versus 5.3% (43/809) by Veritor. Overall, the sensitivity and specificity of Veritor using NAAT as reference standard were 63.1% and 99.7%, respectively (Table 1). A sub-analysis of patients (N=29, average age 52 years, range 5-82 years) with known, recent NAAT confirmed SARS-CoV-2 showed a sensitivity and specificity of 73.9% and 83.3%, respectively (Table 2). For individuals with unknown SARS-CoV-2 status (N=780, average age 45 years, range 0-98), sensitivity and specificity were 57.1% and 99.9%, respectively (Table 2).

**Table 1.** Overall clinical performance of BD Veritor SARS-CoV-2 Antigen test using NAAT as standard reference

| Overall | NAAT-positive | NAAT-negative | Total (N) |
| --- | --- | --- | --- |
| Antigen test positive (N) | 41 | 2 | 43 |
| Antigen negative (N) | 24 | 742 | 766 |
| Total | 65 | 744 | 809 |
| Sensitivity (%) | 63.1% (41/65) | Positive predictive value (%) | 95.3% (41/43) |
| Specificity (%) | 99.7% (742/744) | Negative predictive value (%) | 96.9% (742/766) |

**Table 2.** Clinical performance of BD Veritor SARS-CoV-2 Antigen test using NAAT as standard reference in A) patients with SARS-CoV-2 positive PCR-test within ≤ 72 hrs prior to index test and B) in persons presenting at the Emergency Departments with unknown SARS-CoV-2 status* NPV of a cohort selected for positivity is not relevant.

| A) Patients with recent, NAAT detected SARS-CoV-2 positive status | NAAT-positive | NAAT-negative | Total (N) |
| --- | --- | --- | --- |
| Antigen test positive (N) | 17 | 1 | 18 |
| Antigen negative (N) | 6 | 5 | 11 |
| Total | 23 | 6 | 29 |
| Sensitivity (%) | 73.9% (17/23) | Positive predictive value (%) | 94.4% (17/18) |
| Specificity (%) | 83.3% (5/6) | Negative predictive value (%) | * |
| B) With Unknown SARS-CoV-2 status | NAAT-positive | NAAT-negative | Total (N) |
| Antigen positive (N) | 24 | 1 | 25 |
| Antigen negative (N) | 18 | 737 | 755 |
| Total | 42 | 738 | 780 |
| Sensitivity | 57.1 % (24/42) | Positive predictive value | 96.0 % (24/25) |
| Specificity | 99.9 % (737/738) | Negative predictive value | 97.6 % (737/755) |

Ct analysis was performed on samples with a concurrent RT-PCR result. Higher Ct score on Veritor-to-PCR discordant samples was observed compared to Veritor-to-PCR concordant samples (Figure 2). For discordant samples (N=9), the Ct score average was 25.95± 3.37. In comparison, concordant positive PCR/Veritor samples (N=24) showed an average Ct score of 19.85± 4.94. A two-sided, pooled t-test showed the difference as significant (p=0.003).

**Figure 2.**
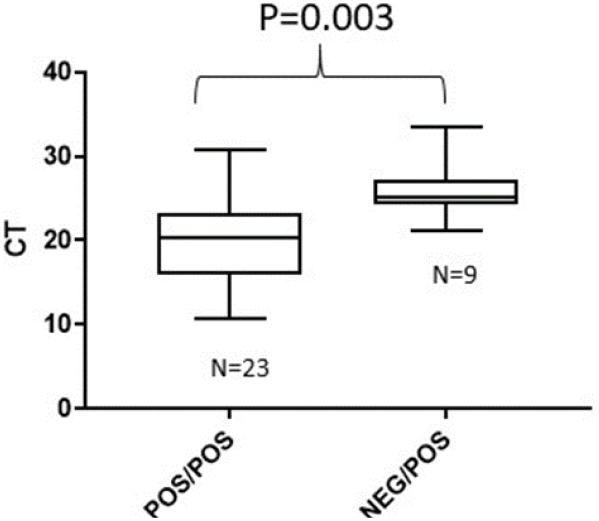
Box plot showing the difference between Veritor-to-RT-PCR concordant and discordant samples

## Discussion

The overall sensitivity of the Veritor test was 63.1%. Though seemingly unimpressive at first glance, the Veritor test performs *at par* with similar antigen test evaluated on the general Danish population^11^ or similar studies with cohorts not dominated by symptomatic persons^6,12-14^. For the group with known, recent NAAT confirmed SARS-CoV-2, the Veritor test provided a sensitivity of 74% (N=29). The sensitivity amongst individuals with unknown SARS-CoV-2 status was 57% (N=780). In comparison, the recent Danish study by Jakobsen *et al*. using the BioSensor antigen test found a sensitivity of 78% for persons with self-reported symptoms, versus 49% amongst people without self-reported symptoms^11^. The specificity of the Veritor test was found to be uniformly high with 99.7% for the overall cohort, and 99.9% in individuals with unknown SARS-CoV-2 status at time of testing.

Amongst those with unknown SARS-CoV-2 status, the Veritor test stands out for the very high PPV of 96% and low Veritor test false-positive rate of 4.0%, which provides confidence that a positive Veritor finding is indeed SARS-CoV-2 positive. This is in contrast to the asymptomatic cohort of the Jakobsen study^11^ where the PPV of the BioSensor antigen test was 72.5%. Finally, the NPV of 97.6% is at par with the majority of antigen test evaluated^6^, with the caveat that high NPV values are driven by the relatively low SARS-CoV-2 prevalence in ours and other’s studies.

Our study cohort is defined by all individuals being in- and outpatients at the Emergency-, Pediatrics- or Infectious Diseases departments and staff undergoing routine testing. The age range is from 5 days old to 98 years. The broad age spectrum contrast with similar studies, most of which only included persons 18 years or older (Table 3), demonstrating the easy operationalization of the Veritor antigen test using a throat collected specimen. The SARS-CoV-2 prevalence by NAAT, overall and amongst persons with unknown SARS-CoV-2 status, was 8.1% and 5.5%, respectively. The prevalence reflects the hospital setting, the enrichment with recently diagnosed, NAAT confirmed SARS-CoV-2 positive patients, and a peak prevalence of SARS-CoV-2 in Denmark at the time.

**Table 3.** Clinical performance characteristics of selected SARS-CoV-2 antigen test systems. N/A: Not applicable, or not reported

| Test | Information source | Study country | Study size (N) | Cohort type | Sampling collection site | Reference standard PCR-test | Sensitivity (%) | Specificity (%) | PPV | NPV |
| --- | --- | --- | --- | --- | --- | --- | --- | --- | --- | --- |
| BD SARS-CoV-2 Veritor | Current study | Denmark | 809 | Complete cohort | Oro-pharyngeal (throat swab) | Roche Flow/ Hologic Pather/ BGI | 63.1% | 99.7% | 95.3% | 96.7% |
|  |  |  | 29 | Recent NAAT test confirmed SARS-CoV-2 positive patients, prevalence 76.6% | Oro-pharyngeal (throat swab) | Roche Flow/ Hologic Pather/ BGI | 73.9 % | 83.3% | 94.4% | N/A |
|  |  |  | 780 | Patients with unknown SARS-CoV-2 Status; Prevalence 5.5% | Oro-pharyngeal (throat swab) | Roche Flow/ Hologic Pather/ BGI | 57.1% | 99.9% | 96 % | 97.6% |
| Q COVID-19 Ag kit (SD Biosensor) | Jakobsen et al.<sup>11</sup> | Denmark | 4811 | General population, (Symptomatic patients: 14.7 %) ≥18 years of age | Throat (PCR) and Nasopharyngeal (Nose) swab (antigen) | Luna Universal Probe One-step RT-qPCR kit (New England Biolab) | Overall 69.7% | Overall 99.5% | Overall 87.0% | Overall 98.5% |
|  |  |  |  |  |  |  | Symptomatic patients: 78.8% | Symptomatic patients: 98.9% | Symptomatic patients: 90.5% | Symptomatic patients: 97.1% |
|  |  |  |  |  |  |  | Asymptomatic patients: 49.2% | Asymptomatic patients: 99.6% | Asymptomatic patients: 72.5% | Asymptomatic patients: 99% |
| BD SARS-CoV-2 Veritor | N. Van der Moeren et al.<sup>17</sup> | Netherlands | 352 | 100 % Symptomatic patients ≥18 years of age | Throat and nose swabs | cobas 6800 (Roche) and m2000 (Abbott) | 80.7% | 100 % | N/A | N/A |
| Q COVID-19 Ag kit (SD Biosensor) | Manufacturer insert | Brazil | 400 | Symptomatic patients (100%) | Nose swab | site-specific RT-PCR method | 88.7% | 97.6% | N/A | N/A |
| Q COVID-19 Ag kit (SD Biosensor) | Manufacturer insert | Germany | 1259 | Symptomatic patients (100%) | Nose swab | site-specific RT-PCR method | 76.6% | 99.3% | N/A | N/A |
| Q COVID-19 Ag kit (SD Biosensor) | Cerutti et al. <sup>12</sup> | Italy | 330 | Symptomatic patients (56%) | Nose swab | Seegene Allplex® 2019 n-CoV Assay, DiaSorin Simplexa®, cobas 6800 Roche® | 70.6 % | 100 % | 100% | 87.4 % |
| Roche SARS-CoV-2 antigen test | Manufacturer insert | Germany | 1039 | Symptomatic patients (84.7%) | Nose and throat | Cobas | 76.6 % | 99.3 % | N/A | N/A |
| Roche SARS-CoV-2 antigen test | Manufacturer insert | Brazil | 392 | Symptomatic patients (98.7%) | Nose | Cobas | 88.7 % | 97.6% | N/A | N/A |
| Panbio COVID-19 Ag Rapid test (Abbott) | Albert et al<sup>13</sup> | Spain | 412 | SARS-CoV-2 PCR prevalence 10.4% | Nose | TaqPath COVID-19 Combo Kit (Thermo Fisher) | 79.6% | 100% | 97.9% | 99% |

Veritor test negative/PCR positive cases had significantly higher Ct scores indicating a lower viral load of the discordant samples compared to the Veritor-to-PCR concordant samples (Figure 2). This is in line with earlier studies of various antigen test system performance^11-13,15-17^. In the respiratory tract, SARS-CoV-2 peak viral load is observed at the time of symptom onset and subsequently declines^18,19^. In a clinical update by Cevik et al.^20^ they conclude that the highest infectiousness potential of SARS-CoV-2 is up to 5 days of symptom onset, but that NAAT tests continue to be positive for a mean of 17 days. Furthermore, Pekosz and colleagues using cell culture demonstrated that the Veritor test had similar performance in detecting replicating SARS-CoV-2 infections as PCR^21^. Nevertheless, as PCR/NAAT is more sensitive than antigen tests or viral cell culture and as human-to-human transmission may occur at even lower levels than observed in cell culture, NAAT remain the reference standard for detection of SARS-CoV-2. From a hospital setting perspective where a fast time-to-result turnaround is required, a positive antigen test should lead to isolation without awaiting confirmatory NAAT testing, whereas a negative antigen test result combined with a SARS-CoV-2 negative clinical evaluation should await NAAT confirmation as the Veritor test cannot exclude SARS-CoV-2 infection.

Comparing antigen test performance across studies is hampered by dissimilarities in study design, cohort characteristics, age, sample collection sites, and reference NAAT test. Table 3 presents antigen test systems in use in Denmark, and it is noteworthy that manufacturer-provided performance data stem from cohorts dominated by very high COVID-19 symptomatic rates resulting in high clinical assay sensitivity estimates not confirmable by real-life, manufacturer-independent studies^11-13^. From a practical perspective, confirming SARS-CoV-2 infection in patients with overt COVID-19 symptoms remains an essential yet minor diagnostic application in absolute numbers. The largest application of antigen testing is screening of predominantly asymptomatic persons as part of efforts to curb dissemination of SARS-CoV-2 infection in society in general and characterized by low prevalence of SARS-CoV-2 infection. Hence the current level of manufacturer-provided documentation and studies focusing on symptomatic cohorts are at best insufficient to guide on antigen test performance in a predominantly non-infected populations undergoing screening by antigen testing.

Our study used an oropharyngeal (throat) collected specimen, which is off-label according to the manufacturers’ specifications and other published studies^11-13,17^. In Denmark, the National Board of Health recommends throat swabs as the preferential sampling site for the detection of SARS-CoV-2. However, the emergence of antigen testing has forced the implementation of nose swabs in Denmark given the lack of data validating antigen tests for throat collected specimens. Combining our study with the Van der Moeren study^17^, the Veritor antigen test could allow the choice of either a throat or nose swab, whatever is preferred by the individual undergoing testing. Moreover, from an operationalization point-of-view, if a patient must undergo an oropharyngeal collected sample for a SARS-CoV-2 NAAT test, then it is possible to do a Veritor antigen test in the same procedure, so that the patient gets two procedures in one test situation.

In a Hospital POCT setting, we found the BD Veritor Antigen test system simple and easily operationalized with positive user experience feed-back. A strength of this study was that the testing was real-life, POCT at multiple departments and the enrollment of samples were representative of the patients seen at the participating departments. All samples were completed immediately on-site, and no frozen or preserved specimens were included. Our POCT setting distinguishes this study from studies using centralized sample testing^6^ or stored samples. The VA unit provides a layer of safe patient identification as the health insurance card is scanned to link patient ID and sample allowing for systematic data reporting. The Veritor test capacity is easily scalable and performs similar whether it is employed using the VA unit or using visual reading of the test cassette^17^. The test buffer supplied neutralizes SARS-CoV-2 virus^22^ which is a safety feature for the staff operating the test system.

All samples were oropharyngeal specimens. This eliminate the potential bias of comparing clinical assay performance between nasopharyngeal, oropharyngeal, nasal, oral or saliva samples. Moreover, access to centralized test databases allowed a detailed record of previous SARS-CoV-2 tests at the individual subject level. A limitation of the study was the comprehensive NAAT technologies for concurrent NAAT testing of reference samples reflecting the real-life hospital situation. This nevertheless limited the power of the Ct-analysis by reducing the number of samples where a Ct score could be retrieved. Regarding Ct score, a general limitation of ours and other’s studies is the interpretation of Ct scores with respect to calling positive cases given the lack of international standardization of PCR Ct values^6^. WHO recently issued a notice of concern regarding interpretation of specimens at or near the limit for PCR detection^23^. A strength of our study was that all positives are called after manual inspection of curves. Yet until joint international standards are agreed, performance evaluations of antigen tests with NAAT as reference standard will benefit NAAT testing in terms of sensitivity.

## Conclusion

In summary, the BD Veritor antigen test for SARS-CoV-2 detection performs well in a hospital POCT setting and could encourage re-thinking whether antigen tests can be used for rapid screening to manage inpatients and outpatients while SARS-CoV-2 status is confirmed by NAAT testing. The very high PPV and NPV in the group of persons with unknown SARS-CoV-2 status jointly demonstrate that the Veritor antigen test represents an analytical improvement for mass-screening compared to the antigen tests recently employed in Denmark. The Veritor antigen tests may serve as a supplement where fast turnaround test results can assist and aide in making clinical decisions regarding SARS-CoV-2 isolation and quarantine but antigen test in general cannot substitute NAAT testing as the primary diagnostic modality for the diagnosis and surveillance of SARS-CoV-2 infection.

## Data Availability

Data are available for external review upon request if provided with relevant Danish Data Protection Agency approvals in concordance with current Danish Law.

## Acknowledgment

We would like to thank the staff of the participating AHH-departments who helped with the collection of samples and data retrieval.

## Financial statement

Sample collection, testing and project management was embedded into the routine of the participating departments. BD Diagnostics, Sparks, MD, donated BD Veritor SARS-CoV-2 antigen tests and Veritor analyzers free-of-charge. The donor had no role in data handling, data presentation, or resulting publication. Marc Arbyn was supported by the Validation of Coronavirus Assays (VALCOR) project (Sciensano, Brussels, Begium).

## REFERENCES

[1] Organization WH: WHO Laboratory testing for coronavirus disease (COVID-19) in suspected human cases: interim guidance. 2020.

[2] Prevention CfDCa: Interim Guidance for Antigen Testing for SARS-CoV-2. 2020.

[3] Kirk JW, Nilsen P: Implementing evidence-based practices in an emergency department: contradictions exposed when prioritising a flow culture. J Clin Nurs 2016, 25:555–65.

[4] Guglielmi G: The explosion of new coronavirus tests that could help to end the pandemic. Nature 2020, 583:506–9.

[5] Guglielmi G: Fast coronavirus tests: what they can and can’t do. Nature 2020, 585:496–8.

[6] Dinnes J, Deeks JJ, Berhane S, Taylor M, Adriano A, Davenport C, Dittrich S, Emperador D, Takwoingi Y, Cunningham J, Beese S, Domen J, Dretzke J, Ferrante di Ruffano L, Harris IM, Price MJ, Taylor-Phillips S, Hooft L, Leeflang MM, McInnes MD, Spijker R, Van den Bruel A, Cochrane C-DTAG: Rapid, point-of-care antigen and molecular-based tests for diagnosis of SARS-CoV-2 infection. Cochrane Database Syst Rev 2021, 3:CD013705.

[7] Peeling RW, Olliaro PL, Boeras DI, Fongwen N: Scaling up COVID-19 rapid antigen tests: promises and challenges. Lancet Infect Dis 2021.

[8] Patterson EI, Prince T, Anderson ER, Casas-Sanchez A, Smith SL, Cansado-Utrilla C, Solomon T, Griffiths MJ, Acosta-Serrano A, Turtle L, Hughes GL: Methods of Inactivation of SARS-CoV-2 for Downstream Biological Assays. J Infect Dis 2020, 222:1462–7.

[9] Corman VM, Eckerle I, Bleicker T, Zaki A, Landt O, Eschbach-Bludau M, van Boheemen S, Gopal R, Ballhause M, Bestebroer TM, Muth D, Muller MA, Drexler JF, Zambon M, Osterhaus AD, Fouchier RM, Drosten C: Detection of a novel human coronavirus by real-time reverse-transcription polymerase chain reaction. Euro Surveill 2012, 17.

[10] Jorgensen RL, Pedersen MS, Chauhan AS, Andreasson LM, Kristiansen GQ, Lisby JG, Rosenstierne MW, Schonning K: An in-well direct lysis method for rapid detection of SARS-CoV-2 by real time RT-PCR in eSwab specimens. J Virol Methods 2021, 289:114062.

[11] Jakobsen KK, Jensen JS, Todsen T, Lippert F, Martel CJ-M, Klokker M, Buchwald Cv: Detection of SARS-CoV-2 infection by rapid antigen test in comparison with RT-PCR in a public setting. MedRxiv early non-peer review pre print 2021.

[12] Cerutti F, Burdino E, Milia MG, Allice T, Gregori G, Bruzzone B, Ghisetti V: Urgent need of rapid tests for SARS CoV-2 antigen detection: Evaluation of the SD-Biosensor antigen test for SARS-CoV-2. J Clin Virol 2020, 132:104654.

[13] Albert E, Torres I, Bueno F, Huntley D, Molla E, Fernandez-Fuentes MA, Martinez M, Poujois S, Forque L, Valdivia A, Solano de la Asuncion C, Ferrer J, Colomina J, Navarro D: Field evaluation of a rapid antigen test (Panbio COVID-19 Ag Rapid Test Device) for COVID-19 diagnosis in primary healthcare centres. Clin Microbiol Infect 2021, 27:472 e7–e10.

[14] Young S, Taylor SN, Cammarata CL, Varnado KG, Roger-Dalbert C, Montano A, Griego-Fullbright C, Burgard C, Fernandez C, Eckert K, Andrews JC, Ren H, Allen J, Ackerman R, Cooper CK: Clinical Evaluation of BD Veritor SARS-CoV-2 Point-of-Care Test Performance Compared to PCR-Based Testing and versus the Sofia 2 SARS Antigen Point-of-Care Test. J Clin Microbiol 2020, 59.

[15] Scohy A, Anantharajah A, Bodeus M, Kabamba-Mukadi B, Verroken A, Rodriguez-Villalobos H: Low performance of rapid antigen detection test as frontline testing for COVID-19 diagnosis. J Clin Virol 2020, 129:104455.

[16] Hirotsu Y, Maejima M, Shibusawa M, Nagakubo Y, Hosaka K, Amemiya K, Sueki H, Hayakawa M, Mochizuki H, Tsutsui T, Kakizaki Y, Miyashita Y, Yagi S, Kojima S, Omata M: Comparison of automated SARS-CoV-2 antigen test for COVID-19 infection with quantitative RT-PCR using 313 nasopharyngeal swabs, including from seven serially followed patients. Int J Infect Dis 2020, 99:397–402.

[17] Moeren NVd, Zwart VF, Lodder EB, Bijllaardt WVd, Esch Hrjmv, Stohr Jjjm, Pot J, Welschen I, Mechelen Pmfv, Pas SD, Kluytmans Jajw: Performance evaluation of a SARS-CoV-2 rapid antigentest: test performance In the community in the Netherlands. medRxiv preprint 72020.

[18] Cevik M, Bamford CGG, Ho A: COVID-19 pandemic-a focused review for clinicians. Clin Microbiol Infect 2020, 26:842–7.

[19] Cevik M, Marcus JL, Buckee C, Smith TC: SARS-CoV-2 transmission dynamics should inform policy. Clin Infect Dis 2020.

[20] Cevik M, Kuppalli K, Kindrachuk J, Peiris M: Virology, transmission, and pathogenesis of SARS-CoV-2. BMJ 2020, 371:m3862.

[21] Pekosz A, Parvu V, Li M, Andrews JC, Manabe YC, Kodsi S, Gary DS, Roger-Dalbert C, Leitch J, Cooper CK: Antigen-Based Testing but Not Real-Time Polymerase Chain Reaction Correlates With Severe Acute Respiratory Syndrome Coronavirus 2 Viral Culture. Clin Infect Dis 2021.

[22] Patterson EI, Prince T, Anderson ER, Casas-Sanchez A, Smith SL, Cansado-Utrilla C, Turtle L, Hughes GL: Methods of inactivation of SARS-CoV-2 for downstream biological assays. bioRxiv 2020.

[23] Organization WH: WHO information notice for IVD users: nucleic acid testing (NAT) technologies that use realtime polymerase chain reaction (RT-PCR) for detection of SARSCoV-2. 2020.

